# Clinician views on best practice community care for people with complex emotional needs and how it can be achieved: a qualitative study

**DOI:** 10.1101/2021.06.17.21259101

**Authors:** Una Foye, Ruth Stuart, Kylee Trevillion, Sian Oram, Dawn Allen, Eva Broeckelmann, Stephen Jefferies, Tamar Jeynes, Mike J Crawford, Paul Moran, Shirley McNicholas, Jo Billings, Oliver Dale, Alan Simpson, Sonia Johnson

## Abstract

**Background:** Service provision for people with complex emotional needs (CEN) is recurrently identified as needing to be transformed: there are serious concerns about quality, accessibility, fragmentation of the service system and the stigma and therapeutic pessimism service users say they encounter. We use the term CEN as a working description to refer to the needs experienced by people who may have been diagnosed with a ‘personality disorder’. Understanding clinician perspectives is vital for service transformation, as their views and experiences shed light on potential barriers to achieving good care, and how these might be overcome. In this study, we aimed to explore these views.

**Methods:** We used a qualitative interview design. A total of fifty participants from a range of professions across specialist and generic community mental health services across England who provide care to people with CEN took part in six focus groups and sixteen one-to-one interviews. We analysed the data using a thematic approach.

**Findings:** Main themes were: 1) Defining Best Practice, 2) Facilitators of Best Care, 3) Barriers to Best Care, 4) Systemic Challenges. Across these themes, staff highlighted in particular the need for care that was person-centred, relational, empathic, and trauma informed. However, major barriers to achieving this are stigmatising attitudes and behaviour towards people with CEN, especially in generic mental health services, lack of development of coherent service systems offering clear long-term pathways and ready access to high quality treatment, and lack of well-developed structures for staff training and support.

**Discussion:** Overall, the findings point towards clinician views as generally congruent with those of service users, reinforcing the need for priorities towards systemwide change to ensure that we can provide the best practice care for these individuals. Particularly prominent is the need to put in place system-wide training and support for clinicians working with CEN, encompassing generic as well as specialist services, and to challenge the stigma still experienced throughout the system.

**Conclusions:** Staff working with this service user group report that delivering best practice care services to be flexible, integrated, and sustainably funded, and for staff to be supported through ongoing training and supervision.

## Introduction

Approximately eight percent of the population have needs that meet the diagnostic criteria for personality disorder [1], a diagnosis that is associated with high rates of co-morbidity [2], premature mortality [3], high levels of service use [4] and high treatment costs [5]. There are serious concerns about the quality and accessibility of services [6] and there is limited evidence to inform service improvement and the implementation of more acceptable, effective and cost-effective models of care [7, 8]. Despite the high levels of distress and reduced quality of life that many people with complex emotional needs (CEN) experience, there is insufficient research to understand the best ways to develop services for people who experience these problems [9]. In considering initiatives to improve the quality and reach of services, co-production with service users will be essential to improve the acceptability and accessibility of services. Understanding clinician perspectives is also vital because they are responsible for delivering care, and therefore their views and experience may directly assist in identifying potential barriers to achieving good care, and how these might be overcome [10].

Many service users report negative consequences as a result of being diagnosed with a ‘personality disorder’, including stigmatisation and exclusion from services [11]. Stigmatising attitudes among mental health professionals and a lack of therapeutic optimism are viewed by service users as significant barriers to the delivery of best practice care [9, 12, 13]. With these issues in mind, in this paper we use the working term CEN to refer to the needs experienced by people who are likely to be diagnosed with a ‘personality disorder’. We use this term CEN in preference to ‘personality disorder’, recognising that many view – and experience – the latter as pejorative and stigmatising [13, 14].

Our recent qualitative meta-synthesis also conducted as part of the NIHR Mental Health Policy Research Unit’s programme to inform NHS policy in this area, summarised literature on clinician perspectives on good practice in community services for people with CEN [15]. These priorities were mainly congruent with those found in studies on service user views [11] and the needs of policy makers [6]. While there was overall agreement regarding what best practice should be, more research is needed to understand the barriers and facilitators to achieving it [15].

This study seeks to fill this gap by exploring staff views on best practice provision of community mental health care for people with CEN and identifying the barriers and facilitators to providing such care. In this study, we focus on the views of staff about the delivery of community mental health care, including both specialist and generic services. This study is part of a programme of research delivered by the NIHR Mental Health Policy Research Unit to inform the design, development and delivery of community mental health services for this population in England [11, 15, 16, 17].

## Methodology

### Design

Qualitative interview study, using individual and focus group interviews.

### Inclusion and Exclusion Criteria

Participants were eligible to take part in the study if they worked in community and outpatient settings at specialist NHS services that provide CEN care, generic (i.e. non-specialist) NHS services that provide generic mental healthcare. Including for people with CEN and those working in social services and the third sector who may be working with some who do not have or do not want access to statutory services. As the focus was on community services, we excluded participants working in forensic, crisis or inpatient settings.

### Recruitment and sampling

Information about the study was circulated via email and social media (e.g. Twitter) to a range of networks, including relevant professional organisations and networks, and workshops/conferences. Interested participants contacted our research team who provided further information about the study, ascertained whether they met the study inclusion criteria, and organised a date for the interview or focus group to take place. Purposive sampling was used to ensure representation of a range of professions and staff who work directly with people with CEN, and of different types of setting, including specialist CEN services, and relevant generic and voluntary sector services. We recruited participants from across England to capture a range of service models and settings.

### Data collection

Data were collected between July 2019 and October 2020. We conducted focus groups and one-to-one semi-structured interviews according to participant preference. Focus groups and interviews were conducted in person or via telephone prior to the COVID-19 pandemic and online via Microsoft TEAMs and Zoom following the introduction of social distancing in March 2020. For remote sessions, participants read and completed online consent forms in advance of the interview, and the researcher reconfirmed consent at the start of the session with recorded verbal consent taken from all participants.

Topic guides were developed with the Lived Experience Advisory Group and service user representatives. These guides were made up of key questions including a) how services currently operate; how services can address the nee ds for service users with CEN; b) how services can support staff to deliver best care; c) what does best practice look like; d) what are the challenges and facilitators to providing this care; e) what are the important outcomes for services and for service users; f) what training and support is needed. Interviews were audio-recorded using either an encrypted digital recorder or the online platform recording option and saved to an encrypted server. Each session lasted between 45 to 90 minutes. Audio recordings of focus groups and interviews were transcribed by an external company. A member of the research team checked the transcripts for accuracy and pseudonymised all participants. All transcripts were allocated a unique ID number and imported to NVivo ProV12 [18] for analysis.

### Analysis

Thematic analysis was undertaken following the phases outlined by Braun and Clarke [19]. Preliminary coding was conducted by UF with members of the wider research team (EB, DA, AS, RS) double coding a selection of interview transcripts. Following initial coding, all themes were reviewed and discussed by the lead author with the wider research team members, who comprised a range of experts by experience and experts by profession. Anonymised data extracts illustrate each theme and key analytic points.

### Ethics

Ethical approval was granted by the Psychiatry, Nursing and Midwifery Research Ethics Subcommittee of King’s College London (reference HR-18/19-10795) prior to data collection with a later amendment to move data collection online in response to COVID-19 restrictions and guidance.

## Findings

### Participant characteristics

A total of 50 participants took part in six focus groups and 16 one-to-one interviews. The sample included 27 women and 23 men. Forty-two participants identified as White, four as Asian, two as Black Caribbean, and two as having other or mixed ethnic background. Participants were recruited from across England, including the Midlands (n=19), North West (n=10), North East (n=6), South West (n=6), London (n=5), and South East (n=4).

Twenty-one participants reported working in specialist ‘personality disorder’ or CEN services, with 29 working in generic community mental health services where individuals with CEN were frequently supported. Most participants were working in NHS services (n=32), with the remaining working in local authority social care settings (n=5) or voluntary/third sector services (n=13).

Participants came from a range of professional backgrounds: psychologists (n=9) and assistant/trainee psychologists (n=5), support workers (n=6), social workers (n=6), peer workers and experts by experience (n=5), psychotherapists and counsellors (5), nurses (n=4), occupational therapists (n=3), psychiatrists (n=3), and commissioner and/ or managers (n=2).

### Themes

Four overarching themes were developed, each with explanatory subthemes outlined in table 1.

**Insert Table 1.** Themes and Subthemes.

| Theme | Subthemes |  |
| --- | --- | --- |
| 1. Defining Best Practice | a.. Best Practice Principles |  |
|  | b. Staff Skills and Characteristics | b.1. Personal Characteristics |
|  |  | b.2. Training, support and Supervision |
| 2. Facilitators of Best Practice | a.. Positive Relationships |  |
|  | b.. Trauma-Informed Care |  |
| 3. Barriers to the Provision of Best Practice | a.. Accessibility and Availability of Services | a.1. Limited and Exclusionary Services |
|  |  | a.2. Limited Access to Specialist Training |
|  | b. Fear of the Risks Associated with Complex Emotional Needs |  |
|  | c. Hopelessness |  |
| 4. Systemic Challenges | a.. Resources and Funding |  |
|  | b.. Stigma and Labelling |  |

#### 1. Defining best practice

There was substantial consensus amongst participants about what constituted best practice for individuals with CEN, with two distinct subthemes: firstly, the principles of best practice care, and secondly, the skills needed to provide best practice care.

##### 1.1. Best practice principles

Participants described several elements of best practice community care for people with CEN (Table 2). Underpinning these principles was the importance of person-centred, individualised care:

**Insert table 2:** Best practice service elements.

| Best practice service elements |
| --- |
| • Person centred<br>• Consistent<br>• Inclusive<br>• Timely and available long term and in times of crisis<br>• Flexible to meet service users' needs<br>• Collaborative across services and sectors<br>• Evidence based<br>• Coproduced with service users<br>• Responding to complexity |

> *“It’s about the ethos of person-centred care… People just need compassion. They need to be loved. As a nurse, that’s what people need to do. That’s what people need to understand, is that a lot of people, they just need people to listen.” (ID 27, female, Expert by Experience working in specialist services)*

Participants also described the importance of responding flexibly to the fluctuating and often complex and intersectional psychosocial needs of service users.

> *“Creation of inclusive pathways and overarching views of the person in multiple systems is needed and needs flexibility to ensure that the pathways bend and flex to the chaos that someone is experiencing.” (ID04, female, Occupational Therapist working in specialist services)*.

Participants described that adapting to the heterogeneous needs and preferences of individual service users necessitated a move away from ‘one-size-fits-all’ models of service delivery towards more flexible care pathways. Many participants also described that responding to the range of service users’ needs (e.g., psychosocial, relational, and medical needs) required collaboration not only between specialist and generic mental health services but also between mental health services and primary care, social services and the third sector.

> *“This just fits with what the Royal College of Psychiatrists is saying: you have a service that spans multiple services and there is something that keeps particular people in mind so that they’re not just disposed of by putting them into a different service or disposed of by not being allowed into one service.” (PT32, male, Occupational Therapist)*.

Coproduction and the involvement of experts by experience was felt by many staff to be a key element that could improve services.

> *“There just needs to be more partnership. It goes back to the fundamental parts of how services should be designed around relationships. There needs to be a relationship between services and this cohort of people so that they are influencing policy and services at commissioner level at the very least. That will ultimately improve people’s lives and save money and make clinicians’ lives easier as well. It just makes everyone’s life easier, I think.” (PT17, male, social worker)*

##### 1.2. Staff skills and characteristics

###### 1.2.1. Personal characteristics

Table 3 shows the personal characteristics identified by staff as important to the provision of effective care for people with CEN.

**Insert Table 3.**
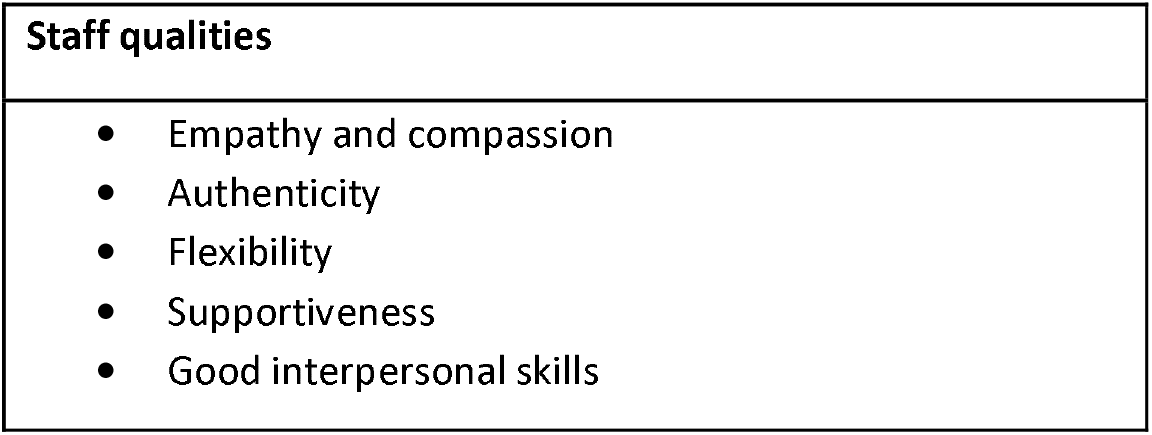
Staff qualities required to provide effective care.

Participants suggested that characteristics such as empathy and compassion enabled staff to see and relate to service users as individuals rather than simply their diagnosis or behaviours, providing care tailored to individuals:

> *“People with characteristics of personality disorder are first and foremost people. So, if you have got the fundamental skills of working with people, then that-The biggest basis for the training is the relational skills. A good friendly manner, a real interest in people, curiosity, thoughtfulness, respect, all that sort of stuff. Honesty, doing what you say to people, which is treating them with respect. All those things.” (ID09, male, psychotherapist working in a CEN service)*

A capacity for reflection and for authenticity in relationships were also seen as essential to the provision of best practice care.

> *“I think just being yourself as well because I think you can’t build an authentic relationship if you are a robot, so maybe I can show a bit more of myself. I think if the problem is building relationships, you can’t build a relationship with someone that’s doing completely textbook and not being human. Just being human in whatever is happening I think is important.” (ID28, female, trainee clinical psychologist)*

###### 1.2.2. Training, support and supervision

Staff skill, which was identified as important to the provision of best practice, could be enhanced through training and support. Training could also help mitigate the impact of compassion fatigue, bad experiences, and exposure to poor practice.

Participants placed high value on training that was coproduced and co-delivered with people with lived experience of complex emotional needs, which, alongside lived experience involvement in service delivery and design, helped provide an “*empathy injection”* to an area often at risk of stigma and preconceptions.

> *“It’s the collaborative training, that is absolutely crucial for practitioners to hear. Because people-We have a very narrow view of each other, we have huge judgments. If we don’t meet people, we know that any kind of attitude change means that you need to understand from the person’s point of view and put yourself in their shoes.” (ID47, male, Clinical Psychologist in generic mental health services)*

Participants also described the need for practitioners in both generic and specialist services to be provided with supervision as well as reflective practice to manage the challenges and emotional demands of providing this care.

> *“That value of supervision and making sure it happens. I could probably go on about that for a while but I think the basic theme is get people thinking about people in a way that promotes empathy and give them enough skills to make them feel confident and competent in working with that client group.” (ID32, male, occupational therapist working in non-specialist services)*

Participants valued the role modelling aspect of supervision and training: the relationships built with supervisors could reflect their aspirations for their therapeutic relationships, for example in providing safety, opportunities for reflection, and the sense of being compassionately held:

> *“I think the training wants to role-model reflection from the trainer, and that they’re emotionally containing of the staff who are coming on the training.” (ID14, female, social worker in a homeless service)*

#### 2. Facilitators of Best Practice

In addition to the skills and characteristics of staff, participants identified two further concepts that facilitate the successful delivery of best practice care: positive relationships and trauma-informed care.

##### 2.1. Positive Relationships

Almost all participants described staff-service user relationships as the most important ingredient of treatment for people with CEN. They explained that many service users had been hurt or rejected by services and that trust in staff and services needed to be established or re-established. Positive staff-service user relationships were important in building trust, and also served to model helpful interpersonal relationships more generally.

> *“We find that people, perhaps, begin to develop a more trusting relationship with us after a period of time when some difficult things have happened. There may have been some incidents, whatever. There may have been some challenges between people in the service and staff team, whatever it might be. Then, we’re six, nine, twelve months down the line, and there’s that sense of trust and actually beginning to open up. We find that’s when it really starts to change because we’ve got a real relationship now.” (ID10, male, social worker in support housing service)*

Empathy, compassion, and authenticity – characteristics reported to be key to the provision of best practice – were described as fundamental to the provision of relational care and to the modelling of supportive but boundaried relationships. Best practice is about holding boundaries and caring enough to have challenging and difficult times together which is what creates the trust and therapeutic bond.

> *“Boundaries are important… it is core to the work that you need to be boundaried as an individual and as an organisation. They [the service user] will violate them and it is the consistency and the way you continue to support then but you do need to have some kind of boundaries and rules but you have to be consistent and explain why so they can learn that abuse isn’t ok but you will keep with them to change and support them.” (PT13, female, clinical psychologist)*

##### 2.2. Addressing Trauma

Participants highlighted the importance of addressing trauma for those with CEN and not focusing solely on presenting symptoms but also engaging with the wider range of difficulties that service users experience and their causes. This included considering how adverse experiences have affected people.

> *“Essentially, we look at things through a trauma lens; “Actually, something happened in your life to lead you to be here today, that’s what we need to figure out.” Nobody wakes up and wants to be in a mental health service for 10 years of their life, nobody does.” (ID42, female, Clinical psychologist in specialist CYP services for CEN)*

Trauma-informed approaches were suggested to provide a way of working with people that was collaborative and empowering. However, participants also conceded that there was a lack of clarity or consensus as to what constitutes a ‘trauma-informed approach’.

> *“I hear a lot of the same things from people across the country. I don’t think it’s an issue in one area, but it is interesting that you say that everyone says, “Oh, I’m trauma-informed,” but it’s not necessarily used right.” (ID26, female, Senior Practitioner in a CEN service)*

This acknowledges that best practice often meant that staff feel they have to do things that service users don’t want at times, but it’s about connecting with service users and providing care with integrity, transparency and compassion.

> *“For trauma-informed, we would want our managers to have the same, to be able to provide the supervision around what’s needed. We’d need ongoing training because we’ve got new staff coming and going all the time.” (PT18, male, counsellor in general mental health services)*

#### 3. Barriers to the provision of best practice

While there is a substantial consensus on what best practice looks like, significant barriers were identified to achieving it. These included the accessibility and availability of services, training and support.

##### 3.1. Accessibility and availability of services

Participants described the limited availability of services for people with CEN and the exclusionary nature of services for those with a ‘personality disorder’ label.

###### 3.1.1. Limited and exclusionary services

In many areas, a combination of restrictive eligibility criteria and long waiting lists made it hard to access appropriate specialist services. Access to generic mental health services was also reported as challenging for those with CEN, with services reportedly excluding service users because of their ‘personality disorder’ diagnosis or restricting care for this reason.

> *“I think services still are set up to exclude people with emotional difficulties… I think any type of ‘personality disorder’ diagnosis or trait is still a diagnosis of exclusion from most places and most services. It firmly does place people in that, kind of, they’re choosing to behave this way and it’s not. It’s not that.” (ID19, female, social worker)*.

> *“It seemed to me that you had to be three quarters on your way to recovery before you could even be accepted by a personality disorder service in the NHS. They were quite rigid. They are also rather risk averse.” (ID09, male, psychotherapist working in a CEN service)*

Exclusions from generic services, e.g., short-term talking therapies such as CBT, was often attributed to service users being “*deemed to be too risky”*. In contrast, in the face of high thresholds and limited services outside generic mental health pathways, participants noted that it often seems as though risk has escalated and the only way into services is via a crisis pathway.

> *“I suppose the other thing is the difficulty of accessing secondary care now*… *I had an emergency myself a while back, a crisis, [it was] two weeks before I could see the psychiatrist. That was the urgent care. That was the, “This is a lady who’s right on the edge.” That is not good service. That is not a good way to treat somebody.” (ID22, female, peer worker in a CEN service)*

Such high thresholds for care were reported to result in an escalation of distress and behavioural disturbance, which services still might not respond to. Seemingly ‘extreme’ behaviours reinforced stigmatising attitudes and a tendency for staff to label service users with CEN as ‘attention-seekers’ or ‘manipulative’.

> *“[some staff say] “Oh, well, they are just attention-seeking”, or, “They have got to take responsibility”, all this kind of stuff… It just sets them up for failure.” (ID08, male, Mental health nurse and psychotherapist in a non-specialist service)*

###### 3.1.2. Limited access to specialist training

Participants reported a lack of specialist training in supporting people with CEN, which they described as a barrier to the provision of quality care by generic mental health services and the third sector.

> *“There is a paucity of training but also a paucity of where they are available. There are quite a lot of courses but if you are in one world and not the other you might not get that information.” (ID24, female, third sector support worker)*

A lack of information about what courses were available, or a lack of support from managers to attend training, exacerbated this problem.

The importance of supervision and reflective practice has been discussed above. Where this was not embedded staff reported feeling less able to engage in the care of people with CEN and to manage risks. Having specialists within teams was also identified as an important way to support staff to provide the best practice care, by providing role modelling opportunities for staff. Without training, supervision and role-modelling, stigmatising attitudes and perceptions were felt to be sustained or born-out of staff burnout and lack of knowledge.

> *“I’ve worked in mental health for the past 20 years. I probably spent the first ten years of my career with very negative views about personality disorder. I think, in retrospect, that’s because nobody ever taught me anything about it, and the only thing that I ever did learn was from people who had also never had any training. So, you know, very, kind of, jaded, burnt-out staff who were very negative*.” *(ID04, female, Occupational Therapist working in specialist services)*.

##### 3.2. Fear of the risks associated with complex emotional needs

Participants described that some staff in generic mental health services were fearful about working with people with CEN because of the severe distress, crisis, recurrent self-harm, and suicidal ideation or intent frequently experienced by this group. Managing such crises and high levels of risk were reported by many staff members as leaving them distressed and feeling an intense sense of powerlessness which was felt to link to therapeutic nihilism, stigma and exclusion.

> *“[Staffs] level of fear gets so high that they think, “I don’t have the resources to do this. They need to be somewhere else.” [this leads to a] buck-passing phenomenon and it’s like when people go from one service to another to another to another.” (ID32, male, occupational therapist)*

Participants suggested that such avoidant and risk-averse behaviours were exacerbated by staff feeling overworked and by a lack of resources and demands to hit targets rather than provide the most effective care for the individual.

> *“When you’ve got a caseload of 50, you’d only need to have 5 people with emotional difficulties on your caseload and your focus is then drawn to them because of the way that they present at services. It would then have a knock-on effect on the outcomes for the other people that were on that person’s caseload, you know, and you’ve got a manager saying, “Well, they’ve done a 12 week DBT programme. Discharge them off now.” Well, that’s not going to work for this person, but you’re stuck by the constraints of the service that you work for and their delivery targets, their KPIs.” (ID19, female, Social Worker working in generic services)*.

##### 3.2. Hopelessness

Several participants working in generic services and in the third sector described finding it challenging to work with service users with CEN, because they felt they could not provide effective help. As a result, some of these staff felt that their role was not about therapeutic support and more about striving to keep people alive.

> *“You make a connection with somebody, you come to care for them. You don’t want them to die.” (ID18, male, counsellor in general mental health services)*

The lack of therapeutic optimism described by some staff was compounded by the lack of resources and services described above. Three participants noted that they had left roles in statutory services as a direct result of these issues.

#### 4. Systemic challenges

##### 4.1. Resources and funding

The under-resourcing of services was reported to directly influence the quality of care for people with CEN, contributing to long waiting lists, delayed discharge, and staff burnout. The importance of sustained investment and funding was repeatedly discussed by participants.

> *“We need money to fund services; without it we have lots of waitlists so there is no seamless care and it makes issues then for other services.” (ID12, female, social worker working with CEN women in homelessness services)*

At the core of discussions about resourcing were that best practice care was relational and took time to do well.

Some staff suggested that the under-resourcing of services for people with CEN reflected a lack of esteem for this group of service users compared to service users with other mental health problems or with physical health problems.

> *“I also think that there’s a lot of stigma about this client group and that, when you are thinking about what your priorities for spending money on are, it’s generally the people who are less popular that get [less] money.” (ID32, male, occupational therapist)*

Some participants argued for the commissioning of integrated specialist CEN and generic mental health services.

> *“I left just disheartened at the fact that actually we’re still working tiered services… we need to skill up a whole workforce and really get away from these tiered pyramid models because it’s everybody’s business.” (ID04, female, Occupational Therapist working in specialist services)*.

Participants also highlighted a need for early intervention services and pathways that allowed service users to progress smoothly through care and to utilise the services that suited their needs.

Important to both joined-up, effective working and to adequate resourcing was having buy-in and commitment from service commissioners and managers. Participants noted that all services are stretched and under-resourced and this has created not only an overworked workforce but also competition across services.

> *“It’s very much to do with culture, and commissioning regimes, and budgets, because that’s the other thing: every budget is so fragmented. So, services are just concerned about their budget and not the overall well-being of that person. How much comes out their budget? “That’s not under my budget. That’s under your budget.” There are all these kinds of structural things that kick in, unfortunately.” (ID18, male, counsellor in general mental health services)*

Several participants reflected that in some cases, services are losing sight of service users’ needs because of budget and resource pressures.

##### 4.2. Stigma and labelling

Most participants highlighted the stigmatisation of individuals with CEN as a systemic problem within healthcare services, with service users excluded from or receiving poorer care from services as a result of their diagnosis. This was particularly felt to be an issue in generic mental health services in which some staff reported having changed the way they employed diagnostic terminology to help service users avoid such exclusionary outcomes.

> *“Every time I assessed someone; I always say ‘Diagnostic uncertainty’ because that meant you were allowed to give them a service. So, you gave them a service, but you didn’t give them a label, then they could come and get a couple of years of help.” (ID16, male, third sector service lead)*

Most participants reported having witnessed stigmatisation and its consequences. Stigmatising attitudes were described as engrained in the culture of many services and attributed to a lack of knowledge about and misunderstanding of CEN as well as negative experiences with service users with ‘personality disorder’ diagnoses.

> *“I think a lot of people just think it is too difficult. It is not as easy to treat as psychosis, depression, all these comparatively easy-to-define presentations. Just I think that they find it easier when it fits into their training and mindset and belief structure. So, coming up against that because it is the dominant-I suppose it is the dominant world view in a lot of services.” (ID17, male, social worker)*

However, many participants also suggested that stigmatising and negative attitudes reflected an attempt by staff to avoid the emotional impact of their work, serving as an unhealthy coping mechanism where support, supervision and training was lacking.

> *“They often are seeing people in crisis, having self-harmed, on in-patient wards*… *I think if I was a nurse, I probably would also say things that aren’t very nice if I didn’t have the containment.” (ID28, male, service lead)*.

Regardless of cause, stigmatisation was understood by participants to be a major system-wide barrier to the provision of best practice care for people with CEN.

#### Discussion

Analysis of interviews with 50 mental health and social care professionals recruited from across England identified best practice principles for providing community care to people with CEN as well as the barriers and facilitators to their implementation. Staff highlighted in particular the need for care that was person-centred, relational and empathic. Importantly, many staff felt that the current provision of care should be informed by an understanding that someone’s current presentation might be shaped by previous experiences of neglect and abuse, often from the very individuals who had been entrusted to care for them earlier in life. However, the ongoing stigmatisation of people with CEN and the lack of staff training and support created barriers to best practice care.

Previous research has emphasised the need for services to focus on the long-term needs of people with CEN, the importance of high quality and consistent therapeutic relationships and of a balanced approach to safety issues, and the need for staff support and joint working across services [15]. Findings from this study add to these recommendations, highlighting in particular the need for services for people with CEN to be more flexible, more integrated, and better resourced.

Most staff considered a “one-size-fits-all” approach inadequate for people with CEN, given the range and fluctuating nature of these needs. However, a lack of flexibility within services and siloed working across services often meant that many needs remained unmet. Addressing the needs of these individuals is likely to require better integration of services and greater flexibility of pathways across both specialist and generic mental health services. This aligns with The Community Mental Health Framework for Adults and Older Adults [20], a mental health policy aimed at greater integration of primary and secondary care and of the voluntary sector in England, and at the development of accessible, straightforward and seamless mental health services. The focus in research on CEN has been almost entirely on testing of specialist psychotherapies, so that very little evidence is available on how to design services and systems of care that deliver high quality and holistic care to meet the long-term needs of people with CEN. Similarly, there is a lack of evidence on whether and how trauma-informed approaches – the use of which was recommended by many participants in this study – contribute to improved outcomes for service users and how they can be best implemented [21].

Although some training programmes have been shown to improve attitudes towards people with a ‘personality disorder’, training alone is unlikely to address what participants described as the systemic stigmatisation of people with CEN [12]. We urgently need evidence about the individual and organisational level factors that contribute to the development and embedding of stigma, including the potential role of fear, risk, and emotional burnout. Also needed are behavioural change perspectives on how to reduce stigma and increase and hopeful and positive attitudes to “personality disorders” as conditions for which there is ample evidence that many people respond to appropriate treatment. While there are barriers and challenging to providing care for those with CEN or a ‘personality disorder diagnosis’, staff need to have understanding that this is a malleable condition that with a lot of improvements see in people who are offered and engage with treatment [22]. It is important for staff to see these outcomes to tackle the hopeless that participants reported within this study.

There is substantial evidence that in England at least, people with CEN continue to experience considerable inequity within services [23]. This study further supports our understanding of this inequality facing individuals with CEN, with staff noting the impact of under-resourcing of services on accessibility of best practice care, as well as the pressures facing staff where training, support and supervision are not resourced for to ensure staff can deliver effective care. This requires service development to not only take into account the needs of service users, but also those of staff, in order to tackle the barriers facing effective care and to integrate elements of best practice such as supervision and reflective practice across services and professional approaches. The findings in this paper also add to our knowledge of what best practice should look like and the often-undervalued elements, such as therapeutic relationships, that are not seen as ‘evidence-based models of care’ but that are imperative to facilitate effective person-centred care.

#### Strengths and limitations

Participants were drawn from across England and from a range of settings and occupations. Topic guides were developed in collaboration with experts by profession and by experience, and these experts also contributed to the analysis and interpretation of data. Some limitations should be noted. Participants were self-selected, and the sample was likely to have included those with an interest and investment in this research, to the detriment of hearing more from staff who work with such service users but were not interested or motivated to take part in a study on this topic. Staff who were less invested in this topic may have different views on, for example, barriers to best practice care, training needs, and the reasons underpinning the stigmatisation of people with CEN.

#### Conclusion

Eighteen years on from the publication of “Personality Disorder: No Longer a Diagnosis of Exclusion” [24], a report which called for the transformation of services for people with a “personality disorder” diagnosis in the NHS many people with CEN continue to be underserved, with barriers to best practice care at the staff, service, and system level. Staff working with this service user group report that delivering best practice care – described as relational, person-centred, and trauma-informed – requires services to be flexible, integrated, and sustainably funded, and for staff across both specialist and generic services to be supported through ongoing training and supervision.

## Data Availability

The data that support the findings of this study are available on request from the corresponding author. The data are not publicly available due to privacy or ethical restrictions.

## Acknowledgements

We would like to acknowledge wider Mental Health Policy Research Unit team, Lived Experience Working Group members, and CEN Steering group who have contributed and supported this work.

## References

[1] Winsper C, Bilgin A, Thompson A, Marwaha S, Chanen AM, Singh SP, et al. The prevalence of personality disorders in the community: a global systematic review and meta-analysis. Br J Psychiatry. 2020 Feb;216(2):69–78.

[2] Shah R, Zanarini MC. Comorbidity of Borderline Personality Disorder. Psychiatric Clinics of North America. 2018 Dec;41(4):583–93.

[3] Fok ML-Y, Stewart R, Hayes RD, Moran P. Predictors of Natural and Unnatural Mortality among Patients with Personality Disorder: Evidence from a Large UK Case Register. PLOS ONE. 2014 Jul 7;9(7):e100979.

[4] Meuldijk D, McCarthy A, Bourke ME, Grenyer BFS. The value of psychological treatment for borderline personality disorder: Systematic review and cost offset analysis of economic evaluations. Schmahl C, editor. PLoS ONE. 2017 Mar 1;12(3):e0171592.

[5] Brettschneider C, Riedel-Heller S, König H-H. A Systematic Review of Economic Evaluations of Treatments for Borderline Personality Disorder. Yukich J, editor. PLoS ONE. 2014 Sep 29;9(9):e107748.

[6] Dale O, Sethi F, Stanton C, Evans S, Barnicot K, Sedgwick R, et al. Personality disorder services in England: findings from a national survey. BJPsych Bulletin. 2017 Oct;41(5):247–53.

[7] Soeteman DI, Hakkaart-van Roijen L, Verheul R, Busschbach JJV. The Economic Burden of Personality Disorders in Mental Health Care. J Clin Psychiatry. 2008 Feb 15;69(2):259–65.

[8] van Asselt ADI, Dirksen CD, Arntz A, Severens JL. The cost of borderline personality disorder: societal cost of illness in BPD-patients. Eur psychiatr. 2007 Sep;22(6):354–61.

[9] Evans S, Sethi F, Dale O, Stanton C, Sedgwick R, Doran M, et al. Personality disorder service provision: a review of the recent literature. Mental Health Review Journal. 2017 Jan 1;22(2):65– 82.

[10] Day NJS, Hunt A, Cortis-Jones L, Grenyer BFS. Clinician attitudes towards borderline personality disorder: A 15-year comparison. Personality and Mental Health. 2018;12(4):309–20.

[11] Sheridan Rains L, Echave A, Rees J, Scott HR, Lever-Taylor B, Broeckelmann E, et al. Service user experiences of community services for Complex Emotional Needs: A qualitative thematic synthesis. medRxiv. 2020 Nov 3;2020.10.30.20222729.

[12] Ring D, Lawn S. Stigma perpetuation at the interface of mental health care: a review to compare patient and clinician perspectives of stigma and borderline personality disorder. Journal of Mental Health. 2019 Mar 12;1–21.

[13] Campbell K, Clarke K-A, Massey D, Lakeman R. Borderline Personality Disorder: To diagnose or not to diagnose? That is the question. International Journal of Mental Health Nursing. 2020;29(5):972–81.

[14] recoveryinthebin A. RITB Position Statement On Borderline Personality Disorder [Internet]. Recovery in the Bin. 2019 [cited 2021 Jun 4]. Available from: https://recoveryinthebin.org/2019/04/03/ritb-position-statement-on-borderline-personality-disorder/

[15] Troup J, Taylor BL, Rains LS, Broeckelmann E, Russell J, Jeynes T, et al. Clinician perspectives on what constitutes good practice in community services for people with Complex Emotional Needs: A qualitative thematic meta-synthesis. medRxiv. 2020 Dec 16;2020.12.15.20248267.

[16] Botham J, Clark A, Steare T, Stuart R, Oram S, Lloyd-Evans B, et al. Community interventions for people with complex emotional needs that meet the criteria for ‘personality disorder’ diagnoses: a systematic review of economic evaluations. medRxiv. 2020 Nov 4;2020.11.03.20225078.

[17] Trevillion K. et al. Service user experiences and perspectives of community services for Complex Emotional Needs: a qualitative study. In preparation.

[18] QSR. NVivo qualitative data analysis software; QSR International Pty Ltd. Version 12, 2018.

[19] Braun V, Clarke V. Special Issue: Coming out in higher education. Lesbian & Gay Psychology Review.2009. 10;1:3–69

[20] NHS England⍰» The community mental health framework for adults and older adults. 2019 [Internet]. [cited 2021 Jun 4]. Available from: https://www.england.nhs.uk/publication/the-community-mental-health-framework-for-adults-and-older-adults/

[21] Sweeney A, Taggart D. (Mis)understanding trauma-informed approaches in mental health. Journal of Mental Health. 2018 Sep 3;27(5):383–7.

[22] Storebø OJ, Stoffers-Winterling JM, Völlm BA, Kongerslev MT, Mattivi JT, Jørgensen MS, et al. Psychological therapies for people with borderline personality disorder. Cochrane Developmental, Psychosocial and Learning Problems Group, editor. Cochrane Database of Systematic Reviews [Internet]. 2020 May 4 [cited 2021 Jun 4]; Available from: http://doi.wiley.com/10.1002/14651858.CD012955.pub2

[23] Anna Freud National Centre for Children and Families, Barnet, Enfield and Haringey NHS Mental Health Trust, The British Association of Social Workers Centre for Mental Health, Mind, Royal College of General Practitioners, Royal College of Nursing and The British Psychological Society. (2017). ‘Shining lights in dark corners of people’s lives’ The Consensus Statement for People with Complex Mental Health Difficulties who are diagnosed with a Personality Disorder. Available at: https://www.mind.org.uk/media-a/4408/consensus-statement-final.pdf [Accessed on 26/04/21].

[24] Personality disorder: No longer a diagnosis of exclusion [Internet]. National Institute of Mental Health; 2003. Report No.: Gateway Ref: 1055. Available from: http://personalitydisorder.org.uk/wp-content/uploads/2015/04/PD-No-longer-a-diagnosis-of-exclusion.pdf

